# Study Protocol for the Pilot Evaluation for SMartphone-adaptable Artificial Intelligence for PRediction and DeTection of Left Ventricular Systolic Dysfunction (The SMART-LV Pilot Study Protocol)

**DOI:** 10.1101/2023.01.30.23285120

**Authors:** Lovedeep Singh Dhingra, Arya Aminorroaya, Veer Sangha, Akshay Khunte, Evangelos K Oikonomou, Bobak J Mortazavi, Robert McNamara, Jeph Herrin, Francis P Wilson, Harlan M Krumholz, Rohan Khera

**Affiliations:** Section of Cardiovascular Medicine, Department of Internal Medicine, Yale University, New Haven, CT, USA; Department of Computer Science, Yale University, New Haven, CT, USA; Department of Computer Science & Engineering, Texas A&M University, College Station, TX; Center for Outcomes Research and Evaluation (CORE), Yale New Haven Hospital, New Haven, CT, USA; Clinical and Translational Research Accelerator, Department of Internal Medicine, Yale University, New Haven, CT, USA; Section of Nephrology, Department of Internal Medicine, Yale University, New Haven, CT, USA; Department of Health Policy and Management, Yale School of Public Health, New Haven, CT, USA; Section of Health Informatics, Department of Biostatistics, Yale School of Public Health, New Haven, CT, USA

## Abstract

**Introduction:** Despite a prevalence of 3-5% among adults, asymptomatic left ventricular systolic dysfunction (LVSD) remains underdiagnosed. There is a critical need for an accurate and widely accessible screening strategy for LVSD, given its association with preventable morbidity and premature mortality. A novel deep learning approach has demonstrated the ability to detect LVSD directly from ECG images, with retrospective validation across multiple institutions. There is a lack of prospective validation. In this pilot study, we evaluate the feasibility of screening and recruiting individuals for prospective echocardiography based on an image-based artificial intelligence (AI)-ECG algorithm applied to the ECG repository at a large academic medical center.

**Research Methods and Analysis:** This is the protocol for a prospective cohort study in outpatient primary care clinics of the Yale New Haven Hospital (YNHH). Adult patients who have undergone a 12-lead ECG without subsequent echocardiogram as a part of routine clinical care within 90 days of the ECG will be identified in the electronic health record (EHR). The AI-ECG model for LVSD will be deployed to YNHH ECG repository to define the probability of LVSD, identifying 5 patients each with high and low probability of LVSD. After discussion with primary care physicians, and subsequent contact by the study team, screened participants will be invited for and undergo an echocardiogram. The study participants and the cardiologists conducting the echocardiograms will be blinded to the results of the AI-ECG screen. The analysis will focus on feasibility metrics: the proportion (i) of all patients undergoing ECGs who have high probability of LVSD without subsequent echocardiogram, (ii) of patients who agree to participate in the study, and (iii) that undergo an echocardiogram. A descriptive exploration of the comparison of the AI-ECG and echocardiogram results will also be reported.

**Ethics and Dissemination:** All patient EHR data required for assessing eligibility and conducting the AI-ECG screening will be accessed through secure servers approved for protected health information. Potential participants will only be contacted after they have discussed the study information with their primary care physician. All participants will be required to provide written informed consent before participation and data will be deidentified prior to analysis. This study protocol has been approved by the Yale Institutional Review Board (Protocol Number: 2000034006) and has been registered at ClinicalTrials.gov (Identifier: NCT05630170). The results of the future validation study will be published in peer-reviewed journals and summaries will be provided to the study participants.

## INTRODUCTION

### Background

Asymptomatic left ventricular systolic dysfunction (LVSD) is highly prevalent in the population, affecting 3-5% of all adults.^1^ However, due to lack of effective screening tools,^1–4^ only 10% of these individuals are diagnosed before the development of symptoms.^1^ Individuals with LVSD have a 5-fold increased risk of developing heart failure and premature mortality, the risk for both of which can be ameliorated with effective treatments.^4–7^ The diagnosis of LVSD has traditionally required cardiac imaging, with limited accessibility. Thus, it is critical to develop an accessible and low-cost screening strategy for LVSD.

The application of deep learning has demonstrated the ability to identify LVSD from 12-lead electrocardiograms (ECGs).^8,9^ However, ECG signal data is rarely available to clinicians and the signal storage formats vary among different machine manufacturers. We have developed a novel deep learning model that is practical for widespread application through its identification of LVSD from ECG images directly, as opposed to raw voltage data used by earlier strategies.^10^ In the form of printed or digital images, ECGs are widely accessible in clinical settings even in low resource areas across the globe. The model performs a pixel-by-pixel assessment of the complete ECG image and can infer complex association in the ECG waveforms and their pixel-level variations at a beat and lead-level to detect signatures of LVSD.^10^ This AI-ECG approach has retrospectively demonstrated high discrimination power across various ECG image formats and calibrations in internal validation (AUROC 0.91, AUPRC 0.55), and external sets of ECG images from outpatient YNHHS clinics (AUROC 0.94, AUPRC 0.77), Cedars Sinai Medical Center in Los Angeles, CA (AUROC 90, AUPRC 0.53), Lake Regional Hospital in Osage Beach, MO (AUROC 0.90, AUPRC 0.88), Memorial Hermann Southeast Hospital in Houston, TX (AUROC 0.91, AUPRC 0.88), Methodist Cardiology Clinic in San Antonio, TX (AUROC 0.90, AUPRC 0.74), and the prospective Brazilian Longitudinal Study of Adult Health (ELSA-Brasil) (AUROC 0.95, AUPRC 0.45). An ECG suggestive of LVSD portended over 27-fold higher odds of LVSD on echocardiogram (OR 27.5, 95% CI, 22.3-33.9 in the held-out set).

However, AI-ECG models have not been prospectively evaluated using protocol-driven echocardiography in contemporary and racially/ethnically diverse populations. This study represents a pilot investigation for the feasibility assessment of a large prospective study for validating AI-ECG image-based LVSD screening.

### Study Objective

Our central hypothesis is that it is feasible to conduct a prospective evaluation of the use of an ECG-image based AI model for the detection of LVSD. The objective of this study is to evaluate the feasibility of using an image-based AI-ECG algorithm in detecting LVSD in a real-world setting.

### Rationale

Given the ubiquitous nature and low of cost of the ECG, and the inaccessible and expensive nature of echocardiograms, the procedure currently used for detection of LVSD, the prospective validation of an AI-ECG based screening strategy could potentially impact a large population of patients. The study design includes a research-indicated echocardiogram following the AI-ECG screen. Echocardiograms are low risk, non-invasive, and without any ionizing radiation. However, early detection of LVSD in asymptomatic patients undergoing echocardiograms could potentially improve outcomes in participants and may reduce or delay progression of the subclinical disease to overt heart failure.^1,5^ Thus, the minimal risk nature of the study intervention along with the significant potential societal benefit provides sound rationale behind the conduction of this pilot study.

## RESEARCH METHODS

### Study Design

This pilot evaluation for the ‘**SM**artphone-adaptable **A**rtificial **I**ntelligence for P**R**ediction and De**T**ection of **L**eft **V**entricular Systolic Dysfunction’ study (the SMART-LV pilot study) follows a prospective cohort study design in the outpatient primary care clinics of the YNHH. The goal of this study is to evaluate the feasibility of recruiting patients and performing an echocardiogram, after pursuing AI-ECG based screening on ECG images. Patients who had undergone a 12-lead ECGs but did not undergo an echocardiogram as a part of routine clinical care within 90 days of the ECG will be retrospectively identified in the EHR. The AI-ECG model for detection of LVSD from ECG images will be deployed to predict the likelihood of missed LVSD that was not evaluated with an echocardiogram, identifying high (≥10%) and low (<10%) probability populations.

For patients screened for eligibility criteria in the EHR, we will discuss the candidacy of the patients with their primary clinician, based on whichever clinician is listed on their Epic chart as their primary care provider. If the clinician decides it is appropriate, the clinician, or their nurse, will contact the patient with information about the study goals and their eligibility for the study visits via an email, my chart message, or a phone call. The clinician or the nurse will explicitly present the patients with the option to opt out of the study. The script used for contacting the patients has been attached as **Appendix 1**. If the patient expresses interest in enrolling in the study or learning more about the study goals, the clinical or the nurse will provide the contact information of the patient to the study team. The research team, through the study coordinator, will call the patient, provide information about the study, and answer any questions the patient may have. Patients expressing interest in participating will be called in for the study visit.

During the study visit, the study coordinators will obtain informed consent for participation in the study. The informed consent form to be used has been attached as **Appendix 2**. Participants will then undergo the echocardiogram, performed by a trained cardiologist. The detailed timeline and study procedures are enlisted in **Table 1**.

**Table 1:** Study Activities and Stages

| Activity | Screening | Study Visit | End of Study |
| --- | --- | --- | --- |
| Implementation of Eligibility Criteria in the EHR | **X** | - | - |
| Identification of Potential Participants with AI-ECG Screening | **X** | - | - |
| Primary Care Physician Approval | **X** | - | - |
| Participant Enrollment | **X** | - | - |
| Confirming Demographic Characteristics | **X** | - | - |
| Informed Consent | - | **X** | - |
| Echocardiogram Procedure | - | **X** | - |
| Participant Compensation | - | **X** | - |
| Adverse Event Reporting | - | - | **X** |
| Feasibility Analysis | - | - | **X** |

### Study Population

Among all patients seeking care at the YNHH, patients who fulfill the following eligibility criteria will be included in the study. To be eligible for inclusion in the study, an individual must meet all of the following criteria:

- Provision of signed and dated informed consent form
- Stated willingness to comply with all study procedures and availability for the duration of the study
- Male or female ≥ 18 years of age at the first visit
- Patients who underwent a 12-lead ECG as a part of their routine clinical care at least 90 days up to 1 year before study recruitment
- Patients who have not opted out of research studies

Any individual who meets any of the following criteria will be excluded from participation in this study:

- Patients who have undergone a prior echocardiogram within 90 days after the 12-lead ECG
- Patients with a prior diagnosis of left ventricular dysfunction, based on a documented low EF in the medical record
- Patients with a prior diagnosis of heart failure as determined by an ICD-10 diagnosis code for heart failure (**Appendix 3**)

### Study Duration

The study is expected to last 6 months, from January 20, 2023 to July 20, 2023, and will use ECGs retrospectively from November 2021 to December 2022 for the AI-ECG model predictions of LVSD. We expect to recruit the participants and schedule research echocardiograms in the first month. The data analysis and study completion report will be prepared in the subsequent months.

### Study Visit

The participants showing interest will be called for study visits for conducting informed consent and research echocardiograms. Each visit will continue for approximately 50 minutes.

The following steps are expected to take place for the research echocardiogram visit:

1. Participants will be consented for participation in the study.
2. The study coordinator will resolve any doubts and clarifications about the study or the procedure.
3. An echocardiogram will be performed, and the results will be interpreted by the cardiologist.
4. The results of the echocardiogram will be shared with the patients. In case of an incidental pathological finding, the report will be added to the patients” EHR and the patients will be referred to their primary care physician or a cardiologist, after consultation with their primary care physician.
5. The participant will receive a pre-paid Visa gift card worth $50 on completion of the study visit.

### End of Study and Follow-up

The study will be completed for each participant after the completion of the echocardiogram, or when they are declared ‘absent for study visit’. Participants who are unable to complete their in-person visits up to 30 days after the initial consent and screening interview will be considered as “absent for study visit”.

The study will be considered complete when the required number of participants have been enrolled, each participant has undergone a research echocardiogram or has been declared ‘absent for study visit’, and the data analysis has been completed.

### Randomization and Blinding

The ECG-image based AI model will be used on all participants undergoing a 12-lead ECG, with an equal number of participants recruited with positive and negative AI-ECG screens. This study does not utilize randomization. The patient participants and the cardiologists conducting the echocardiograms will be blinded to the results of the AI-ECG screen to prevent confirmation bias.

### Outcomes and Analysis

The study will follow the statistical design principles of a study of a diagnostic test with a known gold standard. In this case, the gold-standard test is the echocardiogram. The novel test is the result of our previously developed AI-ECG algorithm. The study does not perform efficacy assessments. It aims to serve as a pilot assessing the feasibility for the prospective evaluation of the performance an image-based AI-ECG algorithm in detecting LVSD in a real-world setting.

The primary analysis is descriptive and will focus on the proportion of patients that screen positive and negative, as well as the proportion of patients who agree to participate in the study, and those that undergo an echocardiogram. The performance of the model will not be assessed in the study. However, to ensure completeness, a descriptive exploration of the comparison of the AI-ECG and echocardiogram results will be reported. These will not be used in the validation data for the algorithm. For this description, LVSD will be defined as an EF<40% based on the results of the echocardiogram and Fischer exact test to compare the proportion of patients with LVSD in the positive AI-ECG screen and negative AI-ECG screen groups

### Sample Size and Study Power

This is a pilot study to examine the feasibility for the prospective evaluation of the performance an image-based AI-ECG algorithm in detecting LVSD in a real-world setting. There is no efficacy evaluation or sample size estimation for the study.

We seek to recruit and perform echocardiograms for a total of 10 patients, 5 patients each in those with high model predicted LVSD, defined as a LVSD probability of ≥10%, and low model-predicted LVSD probability of <10%. We expect a 50% response rate for patients undergoing the study echocardiogram, and will invite 20 patients for the initial screening. If the response rate is lower, more patients will be recruited, and the response rate will be recorded for future studies. We will recruit and conduct echocardiograms for up to 20 patients if all patients respond.

### ETHICS AND DISSEMINATION

The study will be conducted in accordance with the ethical principles that have their origin in the Declaration of Helsinki, Common Rule, Good Clinical Practice (GCP), Institutional Review Board (IRB), and applicable record retention policies. The specific measures taken to ensure ethical conduct and human subject protection are elaborated in sections below.

### Data Storage and Handling

All patient EHR data required for assessing eligibility and conducting the AI-ECG screening will be accessed through The Health Insurance Portability and Accountability Act (1996, HIPAA) compliant YNHH servers, with built-in audit logs and dual factor authentication. The database server is only accessible from within the Yale intranet (or via Virtual Private Network remotely) and additionally requires a separate login username and password. No protected health information will be included in the analysis or publication. All analyses will proceed on this server alone. The linking file will be maintained for potential future linking to national databases. Such linking would not occur in the absence of another IRB-approved protocol. The investigators will permit study-related monitoring, audits, IRB review, and regulatory agency inspections and provide direct access to source data documents.

The data will be maintained on secure, encrypted servers for a minimum of 5 years after the publications of the study findings in a peer-reviewed journal (in such case as there is a need to return to the original source data to validate a finding or respond to a question).

### Human Subject Protection during Enrollment

The healthcare providers will be contacted by the research team for approval to enrol patients in the study. Potential participants to the study will only be contacted after they have discussed the study information with their primary care physician, or their team (**Appendix 1**). This will ensure that the participants have adequate time to discuss all aspects of the research with their primary care physicians before enrolling in the study. The patients will be explicitly provided with the option to not participate in the study by both the primary care physicians and the study coordinators.

### Informed Consent

All participants will be required to provide signed consent before participation. Informed consent will be obtained before the research-indicated echocardiogram. The proposed informed consent form is included in **Appendix 2**.

Informed consent will be administered by the study coordinators to the participants such that sufficient uninterrupted time will be offered for review and clarification of doubts. To ensure participants understand the information presented to them, they will be requested to explain the procedure and its risks in their own words. They will be asked open ended questions to ensure adequate understanding of the study procedures.

### Subject Confidentiality

Subject confidentiality is held in strict trust by the research team. Subject medical record review will be limited to just the elements needed to complete the study. Only authorized HIPAA and GCP-trained study team members will be allowed to extract research data from medical records. Age, sex, and several categories of health information (provider encounters, notes, comorbidities, medication lists, problem lists, family history, allergies, laboratory findings, procedures, immunizations, vital signs, and medical record numbers) and relevant clinical outcomes may be collected on subjects from the EHR. This data will be deidentified prior to analysis.

### Participant Compensation

The participants will be compensated for their time spent participating in the study. We will provide $50 to each participant upon the completion of their study visit. This payment will take the form of a prepaid Visa gift card.

### Management and Reporting of Adverse Events

The use of the AI-ECG model as Software as Medical Device (SaMD) is only for evaluating the ECG images. Hence, the use is associated with minimal risk to the patient. This study design and echocardiogram procedure present minimal risks, and adverse events or medical risks to the participants are not anticipated. In the unlikely event that such events occur, reportable events (which are events that are serious or life-threatening and unanticipated [or anticipated but occurring with a greater frequency than expected] and possibly, probably, or definitely related) or Unanticipated Problems Involving Risks to Subjects or Others (UPIRSOs) that may require a temporary or permanent interruption of study activities will be reported immediately (if possible), followed by a written report within 5 calendar days of the Principal Investigator becoming aware of the event to the IRB (using the appropriate forms from the website) and any appropriate funding and regulatory agencies. The investigator will apprise fellow investigators and study personnel of all UPIRSOs and adverse events that occur during the conduct of this research project during the course of regular study meetings and via email to the fellow investigators. Unrelated events and non-serious, related adverse events will be recorded.

In case of any incidental pathologic finding, participants will be appropriately referred to a primary care physician and/or a cardiologist, in consultation with the primary care physician and any potentially pathologic finding will be added to their electronic health record.

### Institutional Review Board Approval

The study protocol has been reviewed and approved by the Yale IRB (**Protocol Number: 2000034006**).

Any change to the protocol or study team will be implemented after approval from the IRB. The IRB will conduct a continuing review at intervals appropriate to the degree of risk, but not less than once per year. A study closure report will be submitted to the IRB after all research activities have been completed. Other study events (e.g., data breaches, protocol deviations) will also be submitted to the IRB as per the Yale IRB”s policies.

### Study Protocol Registration

The study protocol has been registered at ClinicalTrials.gov (**Identifier: NCT05630170**). Any updates or modifications to the study protocol will be reflected in the public record at ClinicalTrials.gov.

### Plans for Disseminating Study Results

For the future validation study, a manuscript detailing the study design will be submitted for publication and presented at a scientific meeting. Authorship will be determined by mutual agreement and in line with International Committee of Medical Journal Editors authorship requirements.

Summary of the publication will also be made publicly available in the form of blog posts and YouTube videos for the consumption of the public. Summaries will also be communicated to the study participants.

### Patient and Public Involvement

Patients or the general public were not involved in the design of the study protocol.

## Data Availability

All data (excluding protected health information of the participants) produced in the present study will be available upon reasonable request to the authors.

