## Supplementary material for "Study Protocol for the Pilot Evaluation for SMartphone-adaptable Artificial Intelligence for PRediction and DeTection of Left Ventricular Systolic Dysfunction (The SMART-LV Pilot Study Protocol)": Online Supplement

Study Protocol for the Pilot Evaluation for **SM**artphone-adaptable **A**rtificial **I**ntelligence for P**R**ediction and De**T**ection of **L**eft **V**entricular Systolic Dysfunction

**(The SMART-LV Pilot Study Protocol)**

**ONLINE SUPPLEMENT**

**Appendix 1: Script for contacting patients via email, MyChart message, or phone call**

Hello {{Patient Name}},

You are being contacted with information about a new research study being conducted at the Yale New Haven Hospital, for which you are eligible for participation. The study aims to assess the feasibility of using novel methods for identifying structural heart disease from a patient’s ECG using artificial intelligence.

You will be given a $50 gift card for your participation. If you would like to know more about the study, please let us know. We will have the Yale researchers contact you, who will be able to provide more information about the study and answer any questions you may have.

You have the right to decline the offer to participate at any stage, without any changes to your health care services.

Thank you.

**Appendix 2: Informed Consent Form**

### COMPOUND AUTHORIZATION AND CONSENT FOR PARTICIPATION IN A RESEARCH STUDY

**YALE UNIVERSITY SCHOOL OF MEDICINE**

**Study Title:** Pilot Evaluation for **SM**artphone-adaptable **A**rtificial **I**ntelligence for P**R**ediction and De**T**ection of **L**eft **V**entricular Systolic Dysfunction **(The SMART-LV Pilot Study)**

**Principal Investigator (the person who is responsible for this research):**

Rohan Khera, MD, MS

195 Church Street, 6^th^ Floor

New Haven, CT 06510

**Telephone Number**: (xxx) xxx-xxxx

**Research Study Summary:**

- You are being consented to join a research study.
- The purpose of this study is to examine the feasibility of using an artificial intelligence (AI)-driven algorithm for detecting a heart condition called left ventricular systolic dysfunction (LVSD) using your electrocardiogram (ECG). This feasibility study will serve to inform future research on this topic.
- Your ECG was evaluated by the AI model. We are now aiming to compare the results of the AI model’s evaluation with the reports from this echocardiogram, which is the gold standard test to assess for LVSD.
- You will receive a $50 gift card for your participation in the study. This pre-paid Visa gift card will be given upon the completion of your echocardiogram, with disclosure documents included with this card on instructions about the use. We will advise you to read these disclosure documents that accompany the card.
- There is minimal risk from participating in this study. The echocardiogram is a non-invasive test without any radiation. It detects structural heart disease, if present.
- It is possible to have minor discomfort or cool sensation associated with the lubricant gel used during echocardiogram, but there is no risk involved with the gel or the procedure.
- If your echocardiogram reveals any abnormality in the heart that you were previously unaware of, we would refer you to a cardiologist for care for the disease found. This could lead to additional testing and clinic visits if you choose to pursue treatment. This study does not cover the cost of clinical care that you make pursue.
- This study may have not offer direct benefits for you, other than potentially detecting heart disease you were previously unaware of.
- However, the results of this study can benefit the society-at-large as the AI model may be used as a screening tool for LVSD in the future. In this case, many individuals can take advantage of this model for timely diagnosis and treatment for LVSD that improves their outcomes and quality of life.
- Taking part in this study is your choice. You can choose to take part, or you can choose not to take part in this study. You can also change your mind at any time. Whatever choice you make, you will not lose access to your medical care or give up any legal rights or benefits.
- If you are interested in learning more about the study, please continue reading, or have someone read to you, the rest of this document. Take as much time as you need before you make your decision. Ask the study staff questions about anything you do not understand. Once you understand the study, we will ask you if you wish to participate. If so, you will have to sign this form.

**Why is this study being offered to me?**

We are assessing the feasibility of using a machine learning model to predict a type of heart condition, called left-ventricular systolic dysfunction. You are being requested to participate in the study as you are seeking care at a Yale-affiliated clinic, and we have taken approval from your primary care provider before contacting you. We are looking for 20 participants to be part of this pilot study.

**Who is paying for the study?**

This study is not funded, though the research program at the Cardiovascular Data Science (CarDS) Lab receives funding from the National Institutes of Health awarded to the Principal Investigator, Dr. Rohan Khera.

**Who is the Principal Investigator of the Study?**

The Principal Investigator of the study is Dr. Rohan Khera, a general cardiologist at Yale. He is the Director of the Cardiovascular Data Science Lab at Yale, which developed the methodology for the technology being evaluated in the study. Dr. Khera holds a pending patent for the technology, filed through Yale University.

**What is the study about?**

The purpose of this study is to determine the feasibility of using a machine learning model to predict a type of heart condition, called left-ventricular systolic dysfunction.

**What are you asking me to do and how long will it take?**

If you agree to take part in this study, this is what will happen:

You will undergo an echocardiogram or cardiac ultrasound as a part of this study to determine if you have LVSD. An echocardiogram is a graphic outline of your heart’s movement that will take about 20 to 40 minutes. Here is a more detailed explanation of the procedure:

- A trained heart doctor (cardiologist) or sonographer will perform the test. A heart doctor (cardiologist) will interpret the results.
- An instrument called a transducer will be placed on various locations on your chest and upper abdomen and directed toward the heart. This device releases high-frequency sound waves.
- The transducer picks up the echoes of sound waves and transmits them as electrical impulses. The echocardiography machine converts these impulses into moving pictures of the heart. Still pictures are also taken.
- An echocardiogram shows the heart while it is beating. It also shows the heart valves and other structures.

**What are the risks and discomforts of participating?**

Unlike diagnostic procedures that make use of radiation to produce results, there are no major risks involved with echocardiograms. This makes an echocardiogram or a cardiac ultrasound different from other tests like X-rays and CT scans that use small amounts of radiation. It is a non-invasive test, which means you do not have to deal with the pain and complications of an open wound. You may experience minor discomfort associated with the lubricant gel used during echo, but there is no risk involved with the gel.

**How will I know about new risks or important information about the study?**

We will tell you if we learn any new information that could change your mind about taking part in this study.

**How can the study possibly benefit me?**

During this procedure, we may discover a structural heart disease that you didn’t know you have. If this is the case, the results will be added to your electronic health record and you will be referred to a primary care physician or a cardiologist, in consultation with the primary care physician, as appropriate. Outside of this, the study may not benefit you personally. However, it might help individuals in the future timely detect LVSD, before their condition becomes serious.

**How can the study possibly benefit other people?**

The benefits to science and other people may include the clinical application of the AI model in timely detection of LVSD which leads to earlier medical treatment and better outcomes. This may be an important way to identify and treat individuals who may experience heart failure in the future.

**Are there any costs to participation?**

If you take part in this study, you will not have to pay for any services, supplies, study procedures, or care that are provided for this research only (they are NOT part of your routine medical care).

However, if the echocardiogram that has been done for this study indicates a structural heart disease that you were unaware of, you may be referred for additional clinical care. If you choose to pursue care, you or your health insurance must pay for services, supplies, procedures that are part of your routine medical care. You will be responsible for any co-payments required by your insurance.

**Will I be paid for participation?**

The echocardiogram procedure you undergo will be conducted free of cost for you. In addition, you will receive $50 for your participation in the study. We will issue you a pre-paid Visa gift card. We advise you to read the disclosure documents included with your card.

What are my choices if I decide not to take part in this study?

You can choose to not participate in the study. It will not affect your routine clinical care.

**How will you keep my data safe and private?**

We will keep information we collect about you confidential. We will share it with others if you agree to it or when we have to do it because U.S. or State law requires it. For example, we will tell somebody if you we learn that you are hurting a child or an older person.

Your data will be stored in a secure, password-protected server with a study specific ID number. Only study personnel, sponsors, and regulators would have access to this data.

When we publish the results of the research or talk about it in conferences, we will not use your name. If we want to use your name, we would ask you for your permission.

We will also share information about you with other researchers for future research but we will not use your name or other identifiers. We will not ask you for any additional permission.

Your data may be used for future research studies or distributed to another investigator for future research studies without additional informed consent from you. In the event that such data is shared for future studies, all identifiable information will be removed.

**What Information Will You Collect About Me in this Study?**

The information we are asking to use and share is called “Protected Health Information.” It is protected by a federal law called the Privacy Rule of the Health Insurance Portability and Accountability Act (HIPAA). In general, we cannot use or share your health information for research without your permission. If you want, we can give you more information about the Privacy Rule. Also, if you have any questions about the Privacy Rule and your rights, you can speak to Yale Privacy Officer at 203-432-5919.

The specific information about you and your health that we will collect, use, and share includes:

- Research study records
- Medical and laboratory records of echocardiogram done in connection with this study.
- The entire research record and any medical records held by the Yale New Haven Health System created from: 01/01/2015 to 01/01/2025
- Records about phone calls made as part of this research
- Information obtained during this research regarding
  - Echocardiogram reports
  - Results from the medical chart including: Laboratory, x-ray, and other test results
  - Physical exam reports

**How will you use and share my information?**

We will use your information to conduct the study described in this consent form.

We may share your information with:

- The U.S. Department of Health and Human Services (DHHS) agencies
- Representatives from Yale University, the Yale Human Research Protection Program and the Institutional Review Board (the committee that reviews, approves, and monitors research on human participants), who are responsible for ensuring research compliance. These individuals are required to keep all information confidential.
- Health care providers who provide services to you in connection with this study.
- Laboratories and other individuals and organizations that analyze your health information in connection with this study, according to the study plan.
- Principal Investigator of the study
- Co-Investigators and other investigators
- Study Coordinator and Members of the Research Team

We will ensure your information stays private. But, if we share information with people who do not have to follow the Privacy Rule, your information will no longer be protected by the Privacy Rule. Let us know if you have questions about this. However, to better protect your health information, agreements are in place with these individuals and/or companies that require that they keep your information confidential.

**Why must I agree to the information in this document?**

By agreeing, you will allow researchers to use and disclose your information described above for this research study. This is to ensure that the information related to this research is available to all parties who may need it for research purposes. You always have the right to review and copy your health information in your medical record.

**What if I change my mind?**

The authorization to use and disclose your health information collected during your participation in this study will never expire. However, you may withdraw or take away your permission at any time. You may withdraw your permission by telling the study staff or by writing to Rohan Khera, MD, MS, 195 Church Street, 6th Floor, New Haven, CT 06510.

If you withdraw your permission, you will not be able to stay in this study, but the care you get from your doctor outside this study will not change. No new health information identifying you will be gathered after the date you withdraw. Information that has already been collected may still be used and given to others until the end of the research study to ensure the integrity of the study and/or study oversight.

**What if I want to refuse or end participation before the study is over?**

Taking part in this study is your choice. You can choose to take part, or you can choose not to take part in this study. You also can change your mind at any time. Whatever choice you make, you will not lose access to your medical care or give up any legal rights or benefits. Not participating or withdrawing later will not harm your relationship with your own doctors or with this institution.

To withdraw from the study, you can call a member of the research team at +1(203) 747-4429 at any time and tell them that you no longer want to take part.

**What will happen with my data if I stop participating?**

If you stop participating, your data will be fully deidentified. Data collected prior to the time you stop participating would be retained in deidentified form.

**Who should I contact if I have questions?**

Please feel free to ask about anything you don't understand.

If you have questions later or if you have a research-related problem, you can call the Study Coordinator at +1(203) 747-4429.

If you have questions about your rights as a research participant, or you have complaints about this research, you call the Yale Institutional Review Boards at +1(203) 785-4688 or.

**Authorization and Permission**

Your signature below indicates that you have read this consent document and that you agree to be in this study.

| Participant Printed Name |  | Participant Signature |  | Date |
| --- | --- | --- | --- | --- |
| Person Obtaining Consent Printed Name |  | Person Obtaining Consent Signature |  | Date |

**Appendix 3: Heart Failure Diagnosis Codes**

Heart failure will be defined by the following International Classification of Diseases - 10th Revision (ICD-10) codes present in the primary or secondary encounter diagnosis:

'I50' , 'I50.1' , 'I50.2', 'I50.20', 'I50.21', 'I50.22', 'I50.23', 'I50.3', 'I50.30', 'I50.31', 'I50.32', 'I50.33', 'I50.4', 'I50.40', 'I50.41', 'I50.42', 'I50.43', 'I50.8', 'I50.81' , 'I50.810', 'I50.811', 'I50.812', 'I50.813', 'I50.814', 'I50.82', 'I50.83', 'I50.84', 'I50.89', 'I50.9'
